# Examining the evidence for Mendelian randomization homogeneity assumption violation using instrument association with exposure variance

**DOI:** 10.1101/2022.09.12.22279854

**Authors:** Matthew S. Lyon, Louise A. C. Millard, George Davey Smith, Fernando Hartwig, Tom R. Gaunt, Kate Tilling

**Affiliations:** National Institute for Health and Care Research (NIHR) Bristol Biomedical Research Centre, University of Bristol, Oakfield House, Bristol, BS8 2BN, UK; Medical Research Council (MRC) Integrative Epidemiology Unit (IEU), Bristol Medical School (Population Health Sciences), University of Bristol, Oakfield House, Bristol, BS8 2BN, UK; Postgraduate Program in Epidemiology, Federal University of Pelotas, Pelotas, Brazil

**Keywords:** Homogeneity, variance, Mendelian randomization, average causal effect, instrumental variable, interaction

## Abstract

**Background:** Estimation of the average causal effect using instrumental variable (IV) analyses requires homogeneity of instrument-exposure and/or exposure-outcome relationships. Previous research explored the validity of homogeneity assumptions by testing IV-exposure interaction effects using a set of effect modifiers. However, this approach requires that modifiers are known and measured but evidence for interaction may also be observed through IV association with exposure variance without knowledge of the modifier.

**Methods:** We explored the utility of testing for IV-exposure variance effects as evidence against homogeneity through simulation. We also evaluated the approach of removing IVs from Mendelian randomization (MR) analyses that show strong association with exposure variance (hence are likely to have heterogeneous effects). Our methodology was applied to evaluate homogeneity assumptions of LDL, urate and glucose on cardiovascular disease, gout, and type 2 diabetes, respectively.

**Results:** Under simulation, interaction of IV-exposure and exposure-outcome effects by a single modifier led to bias of the estimated average causal effect (ACE) which could be partially assessed by testing for IV-exposure variance effects. Bias of the ACE attenuated after removing instruments with strong exposure variance effects. In applied analyses, we found no strong evidence of bias from the ACE.

**Conclusions:** We find no strong evidence against estimating the ACE for LDL, urate and glucose on cardiovascular disease, gout, and type 2 diabetes. These approaches could be used in future MR analyses to gain improved understanding of the causal estimand.

**Key messages:**

- Homogeneity of the instrument-exposure and/or exposure-outcome effect is necessary to estimate the average causal effect which is important for developing health interventions
- Partial evidence against the homogeneity assumption can be obtained from testing for the instrument-exposure variance effect which may suggest the presence of effect modification
- This evidence can be used in two ways: i) as a falsification approach to determine if the homogeneity assumption may be violated. ii) to remove genetic instruments from Mendelian randomization analyses providing an estimate that is closer to the average causal effect
- After removing instruments with exposure variance effects, the Mendelian randomization effect of LDL, urate and glucose on coronary heart disease, gout, and type 2 diabetes, respectively showed little difference suggesting no strong evidence against the average causal effect

## Introduction

Mendelian randomization (MR) provides causal evidence for an exposure-outcome effect that is less susceptible to confounding and reverse causation than observational associations^1^ and requires three core assumptions (IV1-IV3)^1–3^ which are defined as follows. IV1, the IV is robustly associated with the exposure (relevance assumption)^1–3^. IV2, there are no confounders of the IV-outcome relationship (exchangeability)^1–3^. IV3, the IV only affects the outcome via the exposure (exclusion restriction)^1–3^. IV1-3 assumptions are sufficient to test the sharp null hypothesis that the exposure does not have an effect on the outcome for any individual in the population^2,4^. At least one additional assumption – these are often collectively referred to as IV4 assumptions – is needed to produce a clearly defined causal estimand point estimate and confidence interval^2,4,5^.

Several IV4 assumptions have been proposed and the choice of assumption influences interpretation of the estimate^4,5^. The causal estimand of interest is typically the average causal effect (ACE) which may be estimated under homogeneity of the IV-exposure^8^ and/or exposure-outcome^9^ effect. For a binary exposure, ACE is the average difference in outcome between exposure groups^4,6^. For a continuous exposure, ACE defines the average difference for a one unit increase in exposure^7^.

Recently, the NO Simultaneous Heterogeneity (NOSH) assumption was proposed^4^ which implies ACE can be identified even in the presence of effect modification of either IV association with exposure or exposure-outcome association provided two assumptions are met. The first NOSH assumption states that effect modifiers of the IV-exposure and exposure-outcome effects are independent. The second NOSH assumption states the exposure-outcome relationship is additive linear.

Testing for IV-exposure effect modification may be used as an empirical approach to detect violation of the IV homogeneity assumption^11^. Hypothesised testing of candidate IV-exposure interaction effects to evaluate homogeneity assumptions has been suggested^11^ but this approach may miss unanticipated interaction effects, cannot be used if the modifier is unmeasured, and potentially incurs a large multiple testing burden^12,13^. Alternatively, the presence of effect modification can be identified by testing the association of the IV with exposure variance provided that the exposure is continuous^14–16^. This evidence could be used to evaluate the homogeneity assumptions. Secondly, in a multi-IV setting such as MR^1^, IVs with strong exposure variance effects could be removed from the analysis which may produce a less biased estimate of the ACE.

In this study we explore the utility of testing for IV-exposure variance effects to provide empirical evidence of IV-exposure homogeneity violation using simulation studies. We apply this approach to MR and examples with data from UK Biobank and large GWAS consortia. First, we propose a falsification strategy where IV-exposure variance effects are used to provide evidence against homogeneity. Second, we demonstrate that evidence of IV-exposure variance effects can be used in sensitivity analyses that remove these IVs from MR estimation.

## Methods

### Summary of simulation studies

The following section describes a series of simulation studies (**Table 1**) reported using the aims, data-generating mechanism, estimand, methods, performance (ADEMP) structure^17^. These simulations aim to evaluate the utility of testing IV-exposure variance effects to assess the homogeneity assumption. First, the IV-exposure and exposure-outcome interaction effect size (with a single modifier) was varied to determine the consequences of NOSH assumption one violation on bias of the estimated causal effect (compared to the known ACE) (**Simulation 1**). We also extended this simulation to test the IV-exposure variance effect to determine the conditions under which this information can provide evidence about the bias of the estimated causal effect (compared to the known ACE). Second, we extended this simulation to estimate the relative bias of the estimated causal effect compared with ACE and related the magnitude of this bias to estimates for the strength of the IV-exposure variance effect (**Simulation 2**). Third, we explored the utility of removing IVs with evidence for IV-exposure variance effects from the inverse variance weighted (IVW) analysis on bias of the estimated causal effect (compared to the known ACE) and IVW causal test efficiency (**Simulation 3**).

**Table 1.** Simulation study summary.

| Figure | Aim | Description |
| --- | --- | --- |
| **1** | Causal estimate bias from ACE under NOSH assumption one violation | Simulating NOSH assumption one violation and estimating the causal effect. Determine if the IV-exposure variance test can detect NOSH assumption violation. |
| **2** | Relative causal estimate bias from ACE for increasing interaction effect size and compare this with IV-exposure variance test power | Estimate relative bias of ACE for increasing IV-exposure interaction effect size and fixed exposure-outcome interaction effect. Relate the magnitude of this bias to IV-exposure variance testing. |
| **3** | Determine if removing IVs with interaction effects on exposure can attenuate IVW bias | Estimate the IVW effect using IVs with/without IV-exposure interaction effects. Apply IV-exposure variance test to remove IVs with interaction effects from the MR estimate. |

ACE, average causal effect. NOSH, NO Simultaneous Heterogeneity. IV, instrumental variable.
MR, Mendelian randomization.

### 1. Simulated bias of the average causal effect under NOSH assumption one violation and rejection rate of IV-exposure variance test null hypothesis

#### Aim

To estimate bias of the estimated causal effect compared with ACE under interaction effect of IV-exposure and exposure-outcome by a common modifier (thus violating NOSH assumption one) and relate the bias to IV-exposure variance test null hypothesis rejection rate. The effect of exposure on outcome was additive linear (satisfying NOSH assumption two). The comparison ACE here is the ACE in the population from which the sample was drawn - I.e., with the same distribution of the modifier.

#### Data-generating mechanisms

Data were simulated for n=10,000 independent observations. For the *i*th observation, we simulated a SNP *G*_*i*_ in Hardy-Weinberg equilibrium (HWE) with a minor allele frequency (MAF) of 0.25 scaled to have mean of zero and unit variance. We also simulated a single modifier *U*_*i*_ using the standard Normal distribution. A standard Normal exposure *X*_*i*_ was simulated with SNP main effect *α*_1_ and modifier main effect *α*_2_ each explaining 5% of the exposure variance and SNP-by-modifier *GU*_*i*_ interaction effect *α*_3_ explaining *∈*{0,0.02,0.04,0.06, 0.08,0.1} of exposure variance. The outcome *Y*_*i*_ was simulated to have main effects of the exposure *β*_1_ and modifier *β*_2_ both explaining 5% variance, and 0-10% variance explained by the interaction effect *β*_3_ and residual drawn from the standard Normal distribution. Note that the *GU*_*i*_ and *XU*_*i*_ varied by the same modifier *U*_*i*_ violating the first NOSH assumption. The residual variance for *X*_*i*_ and *Y*_*i*_ was denoted with *E*_1*i*_ and *E*_2*i*_, respectively.

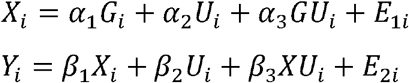

#### Estimand

The IV ACE of *X* on *Y*.

#### Methods

The effect of *G* on *var*(*X*) was tested using the least-absolute deviation regression based Brown-Forsythe test (LAD-BF)^14^. The causal effect of *X* on *Y* was estimated using the Wald ratio.

#### Performance measures

LAD-BF null hypothesis rejection rate was defined as the percentage of repetitions with P < 0.05. Bias of the estimated causal effect compared with ACE was defined as the difference between the Wald ratio estimate and the causal effect used to generate the data i.e., 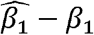. Each configuration of parameters was evaluated using n=500 repetitions.

#### Open-source code

https://github.com/MRCIEU/variance-iv4-violation/blob/master/sim7.R

### 2. Simulated relative bias of the estimated causal effect under NOSH assumption one violation and rejection rate of instrument-exposure variance test null hypothesis

#### Aim

To estimate relative bias of the estimated causal effect compared with ACE from violation of the first NOSH assumption and the IV-exposure variance test null hypothesis rejection rate. This simulation fixed the exposure-outcome interaction effect and varied the IV-exposure interaction effect. Both interactions were varied by the same modifier thus violating the first NOSH assumption. The effect of exposure on outcome was additive linear satisfying NOSH assumption two. This is distinct from **Simulation 2** in that relative bias is used instead of absolute bias. This was to enable lookups of ACE bias and IV-exposure variance test power to guide future studies. Relative bias was chosen so that all estimates were on the same scale. We also included a binary outcome to estimate bias of the estimated causal effect compared with ACE on the log odds ratio to inform future studies.

#### Data-generating mechanisms

Data were simulated for *n*=500, *n*=1000, *n*=2000 and *n*=4000 independent observations for continuous outcomes and *n*=1000, *n*=2000, *n*=4000 and *n*=6000 independent observations for binary outcomes. These sample sizes were chosen to show a range of relative biases across the IV-exposure and exposure-outcome variance explained. For the *i*th observation, we simulated a SNP *G*_*i*_ in HWE with a MAF of 0.25 scaled to have mean of zero and unit variance and standard Normal modifier *U*_*i*_. The standard Normal exposure *X*_*i*_ was simulated with *G*_*i*_ main effect *α*_1_ explaining 1-5% of the variance, *U*_*i*_ main effect *α*_2_ explaining 20% variance, and SNP-by-modifier *GU*_*i*_ interaction effect *α*_3_ 0-2x the size of *α*_1_. Exposures with sample size of *n*=500 and 1% SNP main effect variance explained were not presented as these were susceptible to weak instrument bias (F-statistic less than 10). For the continuous outcome, a standard Normal outcome *Y*_1*i*_ was simulated to have 20% variance explained by *X*_*i*_ and 20% variance explained by *U*_*i*_ and 10% variance explained by the interaction effect *β*_3_ of exposure-by-modifier *XU*_*i*_. For the binary outcome *Y*_2*i*_, the intercept *γ*_0_ was set to logOR (1.1) and the main effects of *X*_*i*_ and *U*_*i*_ were set to logOR (1.1) per 1 SD increase denoted by *γ*_1_ and *γ*_2_. The exposure-by-modifier *XU*_*i*_ interaction effect *γ*_3_ was set to half the size of *γ*_1_. Note that *GU*_*i*_ and *XU*_*i*_ interaction effects vary by *U*_*i*_ violating NOSH assumption one.

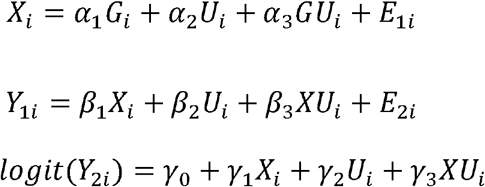

#### Estimand

The IV estimated causal effect of *X* on *Y*_1_ or *Y*_2_.

#### Methods

The effect of *G* on *var*(*X*)was tested using LAD-BF^14^. The causal effect of *X* on *Y*_1_ and *Y*_2_ was estimated using the Wald ratio.

#### Performance measures

LAD-BF test null rejection rate was defined as the percentage of tests with P < 0.05. The relative bias of the estimated causal effect compared with ACE was defined as the simulated Wald ratio divided by the true causal effect for binary (i.e., 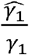) and continuous outcomes (i.e., 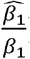), respectively. Each configuration of parameters was evaluated using *n*=500 replications.

#### Open-source code

https://github.com/MRCIEU/variance-iv4-violation/blob/master/sim9.R and https://github.com/MRCIEU/variance-iv4-violation/blob/master/sim11.R

### 3. Simulated average causal effect and IVW test efficiency under NOSH assumption one violation using subsets of instruments ranked by exposure-variance association

#### Aim

To estimate absolute bias of the estimated causal effect compared with ACE and IVW causal test efficiency under violation of the first NOSH assumption for only a subset of IVs. This was performed by including half of the IVs with an interaction effect on exposure in combination with an exposure-outcome interaction effect. We explored the consequences of progressively removing IVs from the IVW analysis on ACE bias and IVW test efficiency by ranking IVs by their association with exposure variance. The effect of exposure on outcome was additive linear holding NOSH assumption two.

#### Data-generating mechanisms

Data were simulated for *n*=100,000 independent observations within each simulated dataset. This large sample size was chosen to obtain precise causal estimates with small numbers of IVs. For the *i*th observation, we simulated six uncorrelated SNPs *G*_*i*_ indexed by *j*, each in HWE and with a MAF of 0.25 scaled to have mean of zero and unit variance. We also simulated a single modifier *U*_*i*_ drawn from the standard Normal distribution. The standard Normal exposure *X*_*i*_ was set to have *G*_*i,j*_ *α*_1,*j*_ main effects drawn from the uniform distribution with sizes of *u*(0.02,0.06) these values were chosen to represent typical genome-wide significant effect sizes. Half of the IVs had an interaction *GU*_*i,j*=1:3_ on *X*_*i*_ with effect size *α*_3,*j*=1:3_ that was set to equal half the size of the main effect *α*_3,*j*=1:3_ × 0.5. The remaining IVs were set to have an interaction *GU*_*i,j*=4:6_ equal to the null *α*_3,*j*=4:6_ = 0. The main effect *α*_2_ of *U*_*i*_ on *X*_*i*_ and the main effects *β*_1_ and *β*_2_ of *X*_*i*_ and *U*_*i*_ on *Y*_*i*_ were set to 20% variance explained. The interaction of *X*_*i*_ and *U*_*i*_ denoted with *XU*_*i*_ explained 10% of the variance *Y*_*i*_. The residual variance of *X*_*i*_ and *Y*_*i*_ were denoted with *E*_1*i*_ *E*_2*i*_, respectively. Note *U*_*i*_ modified the IV-exposure and exposure-outcome relationships violating NOSH assumption one.

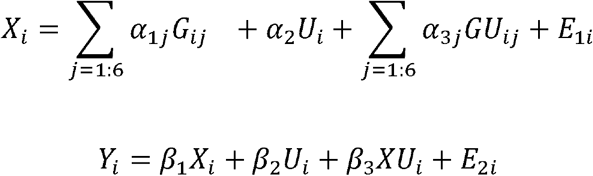

#### Estimand

The IV causal estimate of *X* on *Y*.

#### Methods

The effect of *X* on *Y* was estimated using IVW first using all IVs and then by progressively removing 5%, 10%, 25%, 50%, and 75% of IVs with strongest evidence for IV-exposure variance effect by the LAD-BF test *P*-value^14^. This was compared to the ‘oracle’ method which removed all the IVs with a true (simulated) interaction effect on exposure (i.e., without using the IV-exposure variance test statistic). We anticipated the oracle method to differ from interactions detected by variance only when the IV-exposure interaction effect was incorrectly identified using variance analysis.

#### Performance measures

The exposure-outcome effect. IVW test efficiency, this was estimated using the mean of the IVW standard error between replicates. Each configuration of parameters was evaluated using *n*=500 replications.

#### Open-source code

https://github.com/MRCIEU/variance-iv4-violation/blob/master/sim12.R

### Effect of serum metabolites on disease outcomes

Genetic IVs for mean and variance of randomly sampled urate, glucose and low-density lipoprotein (LDL) cholesterol were extracted from the MRC-IEU OpenGWAS platform^18^ estimated in UK Biobank (**Table S1**). These were applied in a two-sample MR framework to estimate the causal effect of these traits on gout, type 2 diabetes mellitus (T2DM) and coronary heart disease (CHD), respectively using outcome datasets from large consortia (**Table S1**) with non-overlapping samples. The main analysis used all available IVs. Sensitivity analyses were performed by removing 5%, 10%, 25%, 50% and 75% IVs with the strongest IV-exposure variance effects estimated using LAD-BF as potential evidence for NOSH assumption one violation. IV-exposure variance associations were estimated in UK Biobank from our recent study^14^. To ensure any differences in causal estimate between sensitivity analyses was not produced by selecting for weaker IVs (since mean and variance may be correlated), we estimated the mean IV strength (F-statistic; **Supplemental Equation S1 and Equation S2**) for each subset of IVs calculated independently and compared this with the complete set of IVs. This was accomplished by comparing the mean IV-exposure F-statistic for each subset with the mean of all IVs using the Mann-Whitney U test to determine if IVW estimates were affected by differing IV strength.

### Software

All MR estimates were produced using the TwoSampleMR R-package^19^ (v0.5.5). Variant association with trait variance was estimated using the varGWAS R-package^14^ (v1.0.0). All analyses and simulation studies were conducted using R (v3.6.0).

## Results

### Simulated evidence for NOSH assumption one violation using instrument-exposure variance test statistics

The estimated causal effect of continuous exposure on a continuous outcome was unbiased (compared to the ACE used in the data generation process) when either IV-exposure or exposure-outcome interaction effects were null (**Figure 1**). Increasing the IV-exposure and exposure-outcome interaction effect size was associated with increased bias of the estimated causal effect when both relationships were modified by a single variable. This is consistent with violation of the first NOSH assumption. Increasing IV-exposure interaction effect size was also associated with increased strength of IV-exposure variance association (**Figure 1**).

**Figure 1.**
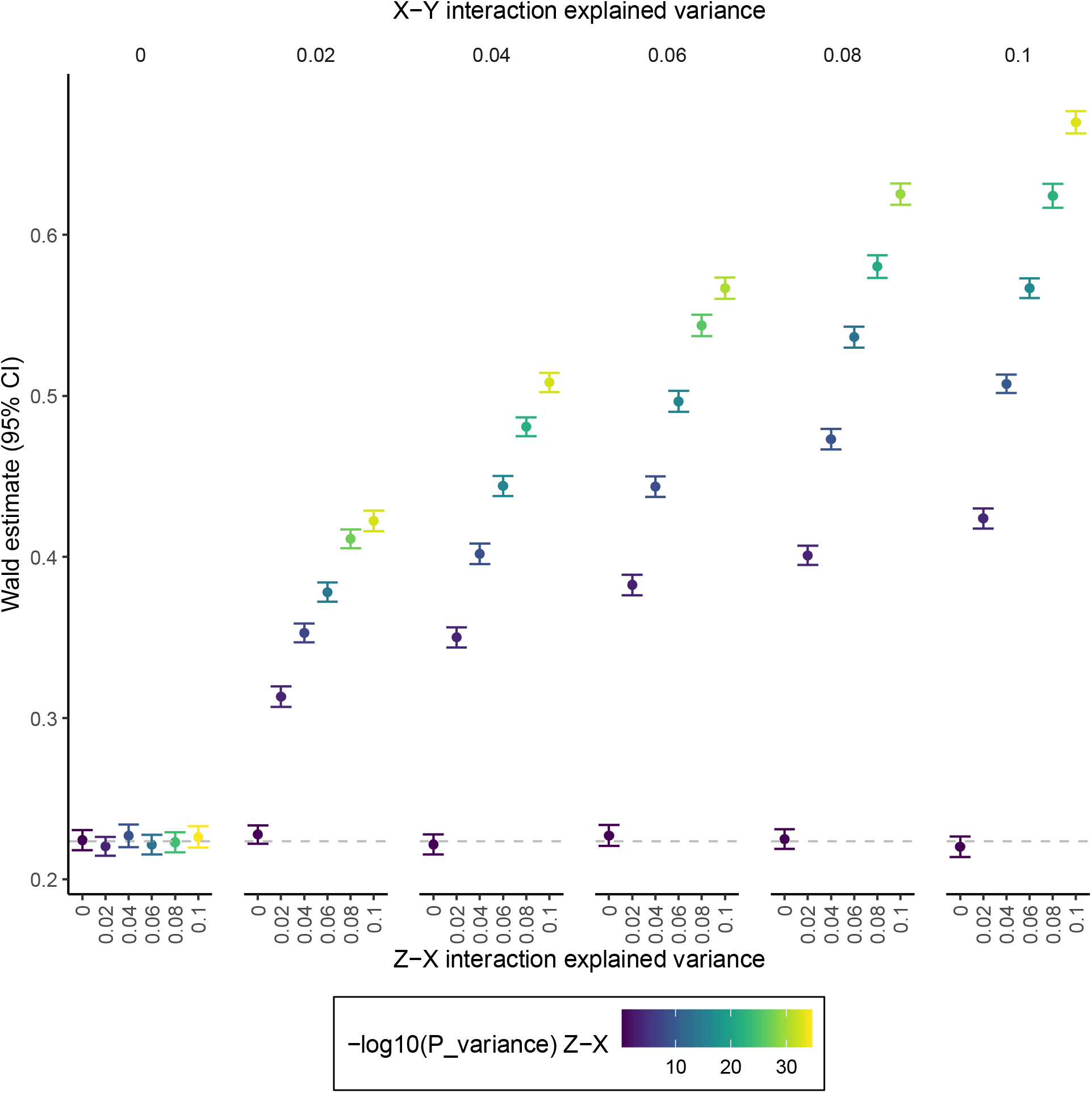
Average causal effect bias under homogeneity assumption violation. Effect of NOSH assumption one violation on causal estimate bias from ACE. Dashed line is expected ACE. Interactions of Z-X and X-Y are modified by the same binary variable (violating NOSH assumption one) but exposure-outcome effect is additive linear (NOSH assumption two holds). Z, instrumental variable. X, exposure. Y, outcome. CI, confidence interval.

Next, we explored the relative bias of the estimated causal effect compared with ACE and related the magnitude of this bias to evidence for IV-exposure variance effects. Relative bias was chosen so that estimates were on the same scale enabling comparisons between parameters and to inform future studies. The strength of IV-exposure variance association was used to indicate potential NOSH assumption one violation. Fixing the continuous exposure-outcome variance explained to 20%, under IV-exposure main and interaction effects of 2% and 1% variance explained respectively, the causal estimate was on average 1.50x the size of the ACE and the IV-exposure variance test rejected the null in 96% of repetitions (95% CI 93%, 97%) with a sample size of n=4000 (**Figure 2**). Fixing the continuous exposure binary outcome effect of 1.1 OR per 1 SD, the magnitude of bias was on average 1.28x times the size of ACE and the IV-exposure variance test null was rejected in 96% of repetitions (95% CI 94%, 98%) given a sample size of n=4000 (**Figure 3**).

**Figure 2.**
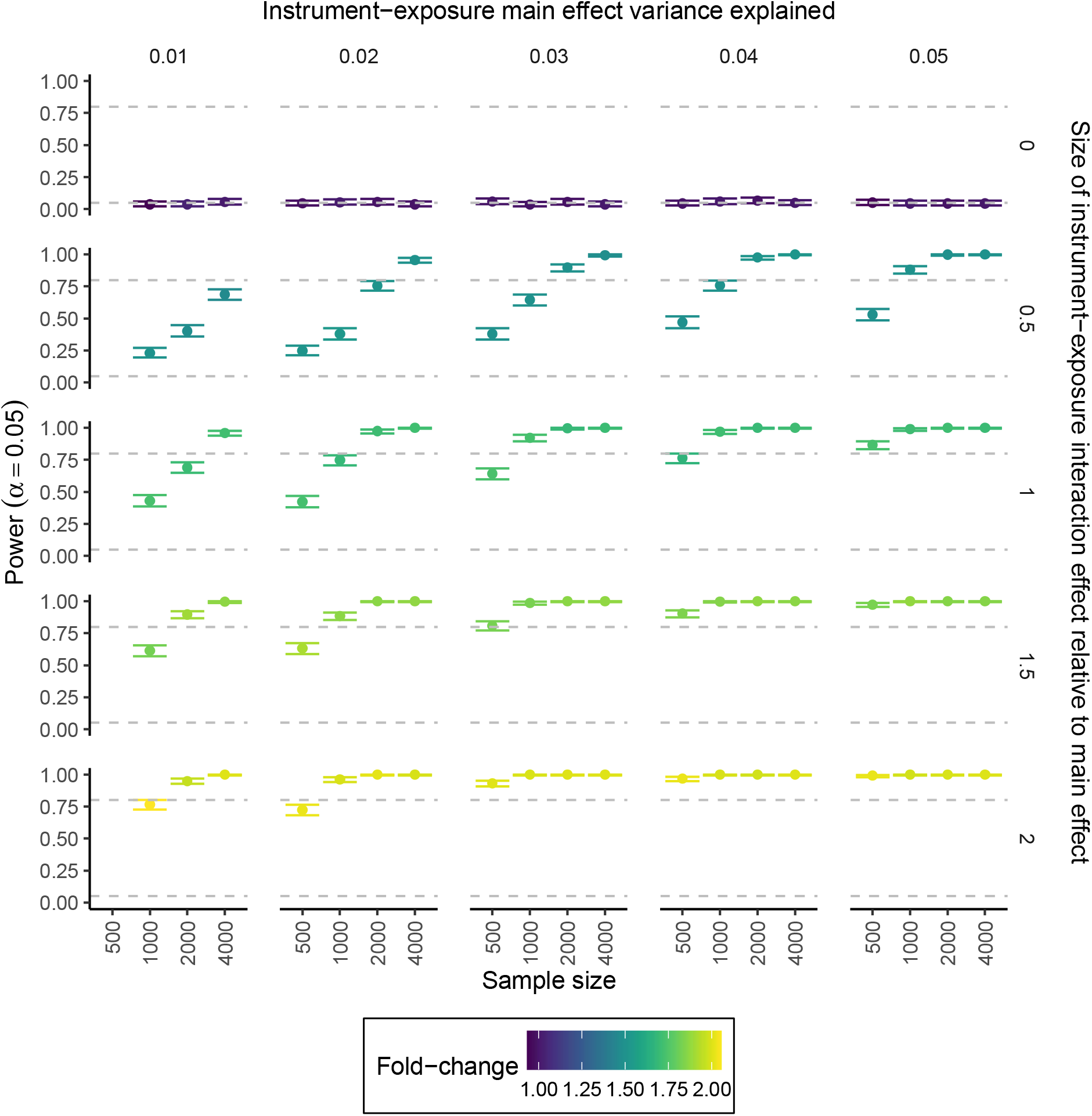
Simulated power to detect instrument-exposure variance effect and relative average causal effect bias under NOSH assumption one violation. Power, proportion of repetitions where the IV-exposure variance test P < 0.05. Fold-change, relative bias of causal estimate from simulated ACE. IV-exposure main and interaction effects were varied. Exposure-outcome main and interaction effects were fixed to 20% and 10% variance explained, respectively. Simulations with n=500 with 1% variance explained by the IV-exposure relationship produced an F-statistic less than 10 and were not shown. Dashed lines represent 5% and 80% power.

**Figure 3.**
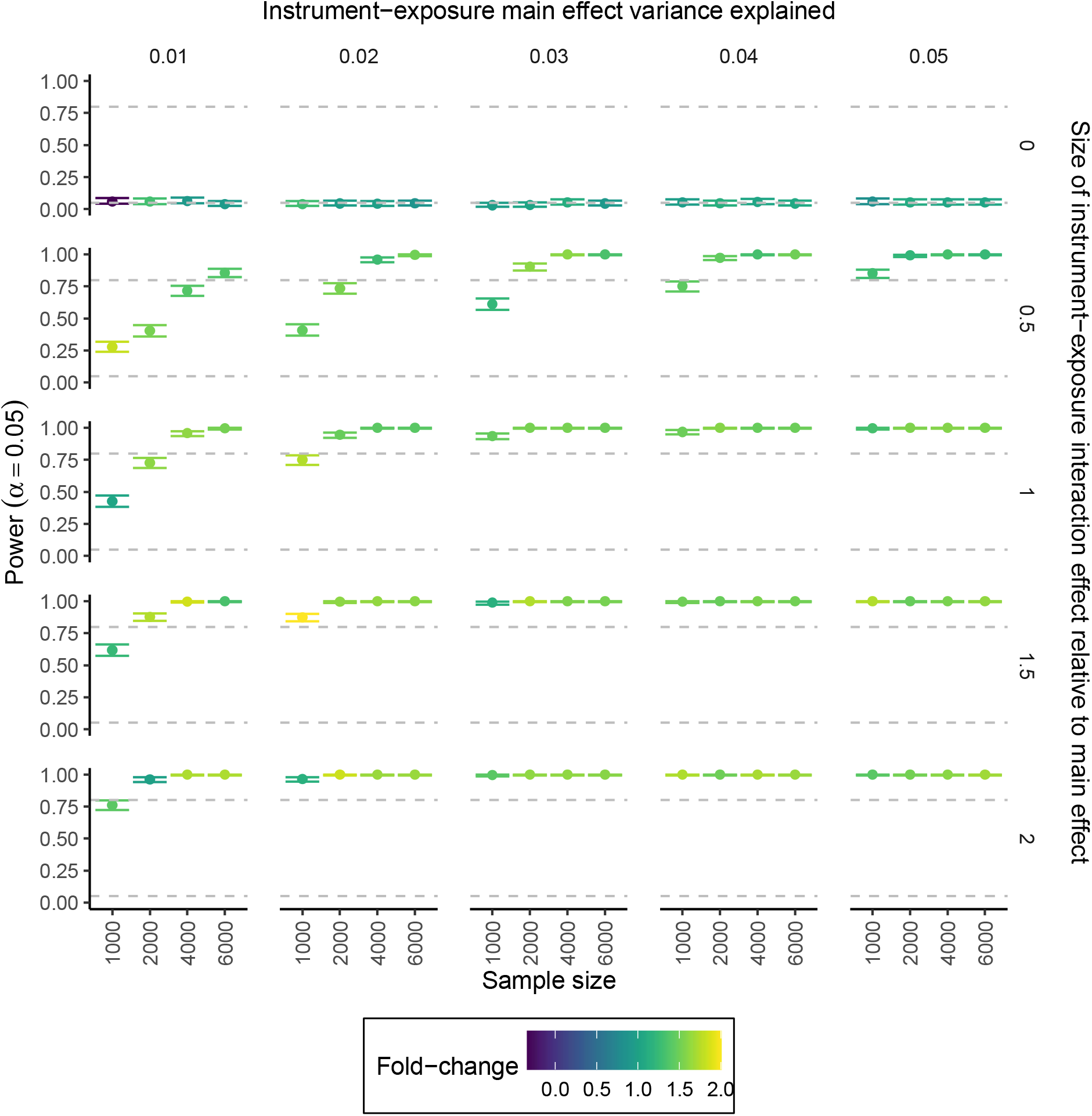
Power to detect instrument-exposure variance effect and relative average causal effect bias of binary outcomes from NOSH assumption one violation. Power, proportion of repetitions where the IV-exposure variance test P < 0.05. Fold-change, relative bias of causal estimated compared with simulated ACE on the log-odds scale. IV-exposure main and interaction effects were varied. Exposure-outcome main and interaction effects were fixed to 1.1 OR. Dashed lines represent 5% and 80% power.

### Simulated effects on average causal effect bias and statistical efficiency of removing instruments by strength of association with exposure variance

Under simulation, we explored the consequences on bias of the estimated causal effect compared with ACE and statistical efficiency of IVW by removing IVs from analysis which were associated with exposure variance (**Figure 4**). Half of the IVs were simulated to have an interaction effect on the exposure and the exposure was simulated to have an interaction effect on the outcome. All interaction effects had the same single modifier, violating NOSH assumption one. Instruments were progressively removed from the IVW analysis using IV-exposure variance test strength by *P*-value estimated with LAD-BF (**Figure 4**). IVW estimates were less biased when IVs with exposure variance effects were removed but this also led to larger IVW standard errors. For example, using all the IVs including 50% simulated with an interaction effect on the exposure, the average causal estimate was 0.53 SD (95% CI of 0.52-0.54) per 1 SD exposure in contrast to the simulated effect of 0.447 SD. Removing the top 50% of IV-exposure variance effects produced an average causal estimate of 0.45 SD (95% CI 0.44-0.46) in line with the simulated effect. This estimate was also consistent with the oracle method which removed IVs simulated to have non-zero exposure interaction effect (0.45 SD [95% CI 0.44-0.47]). The oracle results could differ if the IV-exposure variance test incorrectly removed/retained SNPs with interaction effects from the model. However, fewer IVs led to reduced statistical efficiency of IVW (**Figure 4**). When all IVs were employed the average standard error for the causal effect estimate was 0.05 (95% CI 0.04-0.05) but this increased to 0.07 (95% CI 0.07-0.07) after removing 75% of IVs with variance effects.

**Figure 4.**
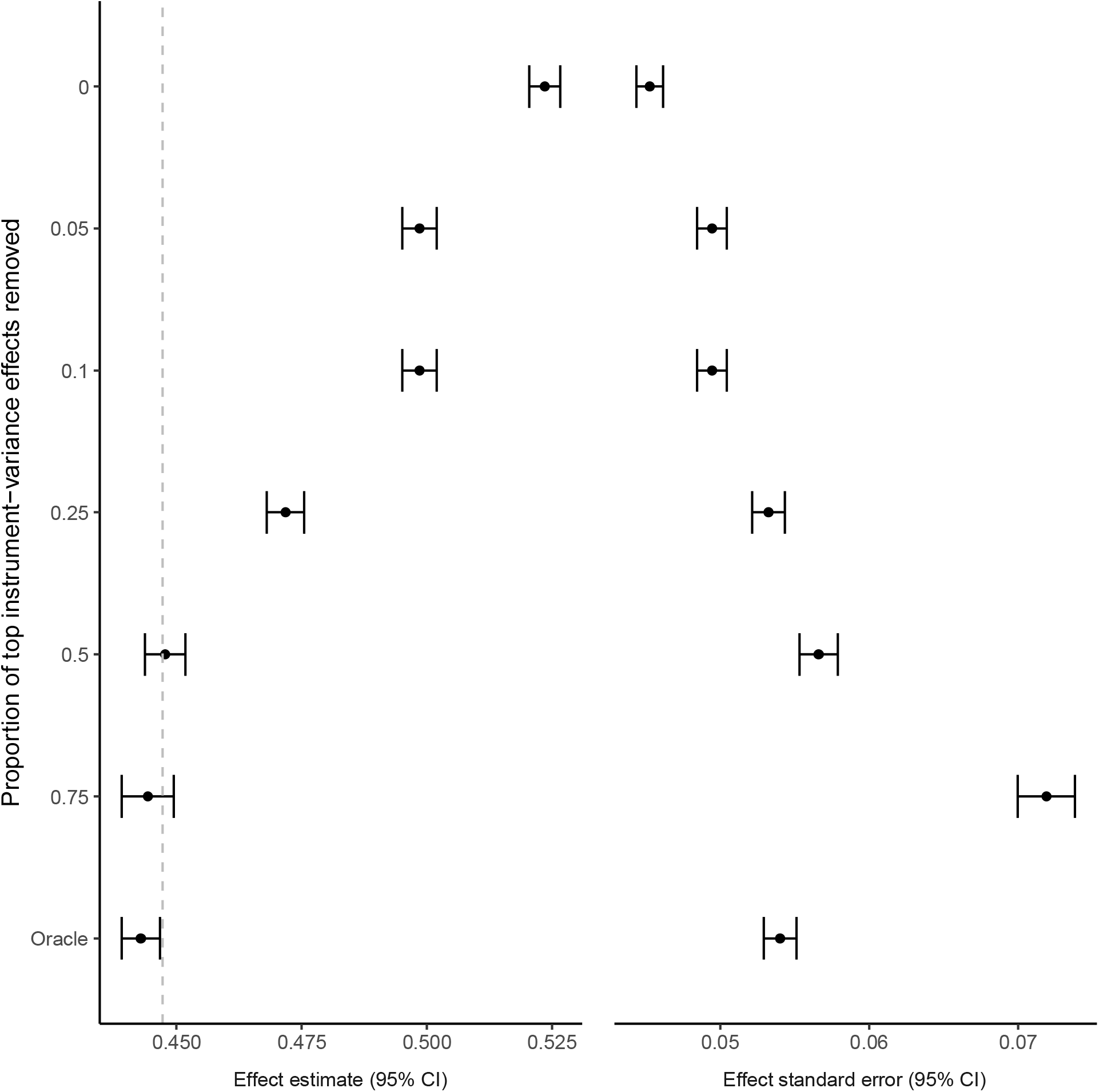
Simulated effect of removing instruments with exposure variance effects on average causal effect bias and statistical efficiency. IVW effect of simulated exposure on outcome using subsets of IVs by IV-exposure variance effect strength and mean of IVW effect standard error. The effect estimate subplot provides the mean causal estimate for each analysis. The effect standard error subplot shows the mean standard error for each analysis. SD, standard deviation. CI, confidence interval. SE, mean IVW standard error of 500 replicates.

### Applied analyses of metabolite concentration on disease outcomes

IVW effect estimates of serum metabolites on disease outcomes were produced using IVs stratified by IV-exposure variance effect strength (**Figure 5**). Starting with the complete set of IVs, the per SD exposure causal effect estimates were: 3.33 OR (95% CI 1.44-7.66) for glucose effect on T2DM, 1.78 OR (95% CI 1.52-2.09) LDL on CHD and 3.26 OR (95% CI 3.00-3.54) urate on gout. As IVs were removed from the analysis, causal estimates attenuated towards the null. This was most extreme for LDL-CHD which reversed sign giving evidence for a protective effect of 0.58 OR (95% CI 0.29-1.20) after removing the top 75% of IVs ranked by IV-exposure variance strength. However, this group of IVs was also weaker (F-statistic mean of 40.56 [95% CI 35.50-46.73]) compared with the full set (F-statistic mean of 71.62 [95% CI 59.69-88.91]) suggesting weaker IVs may be responsible for biasing these effects. Where there was little evidence of weaker IVs, we detected little attenuation of point estimates compared with the full set of IVs and overlapping confidence intervals suggested no strong difference. For example, the effect of LDL-CHD excluding the top 10% of IV-variance effects gave a causal estimate of 1.44 OR (95% CI 1.22-1.84) and removing the top 75% of IVs led to estimates of 3.17 OR (95% CI 1.71-5.87) and 2.56 OR (95% CI 2.10-3.14) for glucose on T2DM and urate on gout, respectively. As these estimates were consistent with the full set of IVs, there was no strong evidence for bias due to violation of the homogeneity assumption. However, it is also possible that IVs with the weakest evidence for exposure variance effect have an interaction effect on the exposure, but the LAD-BF test power was too low to detect an effect.

**Figure 5.**
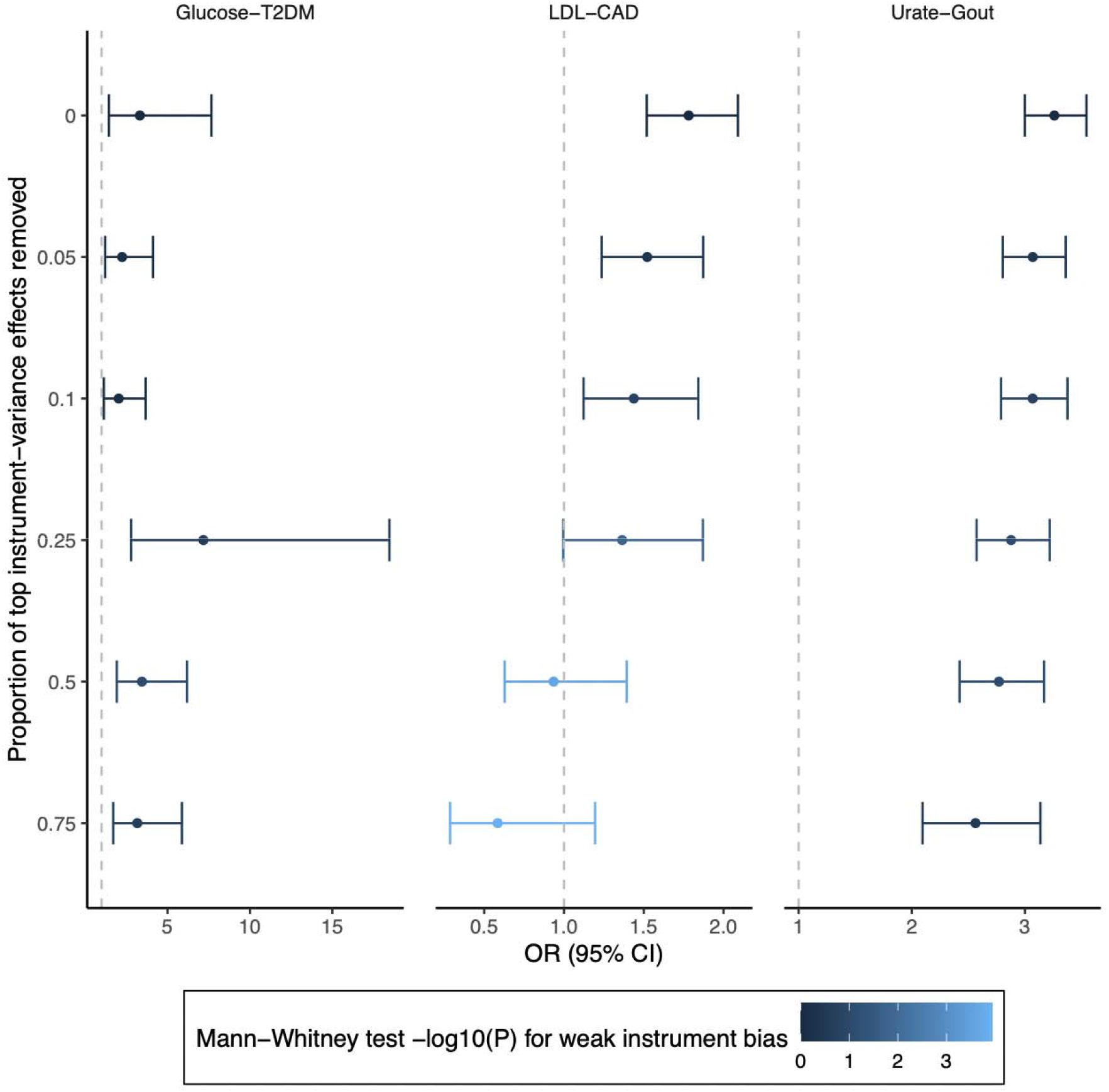
MR sensitivity analysis removing instruments with exposure variance effects. Inverse-variance weighted effect of serum metabolite concentration on binary outcomes. OR, odds ratio. CI, confidence interval. LDL, low-density lipoprotein. CAD, coronary artery disease. T2DM, type II diabetes mellitus. Mann-Whitney U test is for the comparison of IV-exposure F-statistic mean effect for all IVs vs the subset as a measure for conditioning on IV strength.

## Discussion

Under NOSH the IV analysis estimates the ACE^4^, but the two NOSH assumptions cannot be completely assessed using the observed data. Here we demonstrated via simulation the potential in testing of IV-exposure variance effects for continuous exposures as an empirical approach to partially evaluate NOSH assumption one. Testing of IV-exposure variance effects cannot prove that NOSH assumption one holds as power may be too low detect a variance effect of IV-exposure where a true interaction exists. Further, even if there is an interaction effect of IV-exposure then NOSH assumption one is only violated if the exposure-outcome effect is modified by this same variable. Secondly, we show IV-exposure variance evidence could be used to mitigate bias from the causal estimate by removing IVs with strong exposure variance effects. This methodology was applied to GWAS summary statistics generated in UK Biobank and non-overlapping large consortia.

Simulations showed that the approach was well powered to detect bias from ACE using IV-exposure variance effects when using the large sample sizes which are now readily available from biobanks. However, IV-exposure variance association evidence cannot specifically identify NOSH violation. The first NOSH assumption requires effect modification of the exposure-outcome and IV-exposure relationships to be independent^4^ but this scenario is not specifically evaluated using measures of IV effect on exposure variance. Lack of an IV-exposure variance effect could suggest that the IV estimand estimates ACE subject to sufficient power to detect an effect. Identification of IV-exposure variance effects could enable follow up studies to identify the precise exposure-modifier interaction effect^13,14,16^. This could be useful to consider if this variable also modifies the exposure-outcome effect which would then imply NOSH assumption one is violated. Conversely, NOSH violation may occur without IV-exposure variance association through non-linearity of the exposure-outcome relationship (NOSH assumption two)^4^.

This approach of testing for violation of homogeneity is similar in principle with Brookhart et *al*^11^ who suggest testing for IV-exposure interaction effects. However, Brookhart et al require effect modifiers to be hypothesised and measured while the approach outlined here is based on IV-exposure variance association and does not.

We explored the utility of eliminating IVs based on their association with exposure variance to determine if bias of the IV estimate diminished through simulation studies. We observed reduced bias of the causal estimate from ACE but also wider confidence intervals which is anticipated because fewer IVs were included in the causal model. Nevertheless, this approach could be useful as a sensitivity analysis to determine if the main analysis (i.e., using all IVs) produces an estimate strongly different from a subset of IVs with the least exposure variance association. While we used the strength of association between IV and exposure variance, future studies could explore the magnitude of effect (i.e., using the variance effect size) but with LAD-BF two coefficients are produced for each allele copy and are difficult to rank.

This approach was applied to evaluate the effects of serum LDL, glucose, and urate on CHD, T2DM, and gout, respectively. IVW estimates did not robustly differ after removing IVs with IV-exposure variance effects for glucose-T2DM and urate-gout suggesting no strong evidence of causal estimate bias from ACE. Meanwhile, the effect directionally of LDL-CHD was reversed when the top IV-exposure variance effects were removed. Two possibilities may explain this finding.

First, IVs may be acting via distinct causal pathways, for example a previous study found a cluster of IVs for BMI that had protective effects on cardiovascular disease^20^ whereas the remaining IVs were associated with adverse effects. Furthermore, evidence for IV-exposure variance association could arise under horizontal pleiotropy^21^, for example if the IV is acting on the exposure through several pathways and some of these are influenced by effect modification. This is in contrast with, for example, an IV within the cis region of a protein coding gene which is less likely to be affected by horizontal pleiotropy^22^.

Second, weak instrument bias may be introduced by conditioning on weaker IV-exposure variance effects (since both mean and variance effects can be correlated^23,24^). While the mean F-statistic was above the rule of thumb of ten^25^ these estimates could be inflated due to chance which is known to introduce bias to causal estimates^25^. In the two-sample MR framework weak instrument bias causes estimates to attenuate towards the null in contrast with two-stage least squares approach which introduces bias towards the observational association^26^. One way to avoid this is to use two-sample MR with second-order weights which incorporate imprecision in the IV-exposure relationship into the causal estimate^27^.

Testing for IV-exposure variance effects could be applied to future MR studies as a sensitivity analysis to determine if effects may be biased from ACE. This approach could also be developed further using meta-regression^28^ techniques to add less weight to IVs that have strong variance effects rather than simply removing these IVs and may preserve statistical efficiency.

However, this work also has some limitations. First, non-normality of the exposure may reduce power and increase type I error rate of the IV-exposure variance association^16^. Second, removing IVs from IV analyses (e.g., by the association of IV with exposure mean or variance) may bias the causal estimate standard errors^21^ leading to type I error rate inflation. This could also lead to conditioning on IVs that exhibit horizontal pleiotropy introducing bias into the estimate^21^.

Through this work we evaluated the strength of the IV-exposure variance association as an empirical approach to evaluate IV-exposure homogeneity assumptions which may be used in falsification studies to determine if the causal estimated is biased ACE, and for sensitivity analyses to provide evidence against ACE as the estimated estimand. We applied these methods to evaluate the effects of LDL-CHD, urate-gout and glucose-T2DM but found no strong evidence of bias from ACE. This approach could be applied to future IV studies to improve interpretability of IV estimates.

## Data Availability

GWAS summary statistics are available from https://gwas.mrcieu.ac.uk/.

## Data and open-source code availability

R-code for simulations, Mendelian Randomization studies and UK Biobank analyses are available from https://github.com/MRCIEU/variance-iv4-violation. UK Biobank data are available from https://www.ukbiobank.ac.uk.

## Funding

This study was funded by the NIHR Biomedical Research Centre at University Hospitals Bristol and Weston NHS Foundation Trust and the University of Bristol. The views expressed are those of the author(s) and not necessarily those of the NIHR or the Department of Health and Social Care. This work was also funded by the UK Medical Research Council as part of the MRC Integrative Epidemiology Unit (MC_UU_00011/1, MC_UU_00011/3 and MC_UU_00011/4). L.A.C.M is funded by a University of Bristol Vice-Chancellor’s fellowship.

### Ethical approval

Ethical approval was not required for this study. All analyses were conducted with simulated data or freely available GWAS summary statistics.

