## Supplemental Text for "Examining the evidence for Mendelian randomization homogeneity assumption violation using instrument association with exposure variance"

**Supplemental information**

*Estimation of SNP variance explained and F-statistic from GWAS summary data*

The F-statistic was used as a measure of instrument strength and was estimated from GWAS summary statistics (**Supplemental** **Equation 1**) as previously described^1^.

**Equation S1. Estimation of the F-statistic from R^2^**

$$F=R^{2} \times(n-1-k)/(\left( 1-R^{2} \right)\times k)$$

Where $n$ is the sample size, $k$ is the number of single nucleotide polymorphisms (SNPs) included in the model ($k=1$ in this analysis), and $R^{2}$ is the variance explained by the SNP which may be estimated using **Supplemental Equation** **2**.

**Equation S2. Estimation of R^2^ from GWAS summary statistics**

$$R^{2}=2\beta^{2}\times\left( MAF \right)\times(1-MAF)/(2\beta^{2}\times\left( MAF \right)\times\left( 1-MAF \right)+\left( se\left( \beta\right) \right)^{2}\times2n\times MAF\times\left( 1-MAF \right))$$

Where $\beta$ is the average effect of the SNP on the trait, $MAF$ is the minor allele frequency and $se$ is the standard error of $\beta$.

**Table S1. Sources and characteristics of GWAS summary statistics**

| **Outcome** | **Source** | **Sex** | **Population** | **N** | **Cases** | **Controls** |
| --- | --- | --- | --- | --- | --- | --- |
| Type 2 diabetes | DIAGRAMplusMetabochip^2^ | Males and females | Mostly European | 149,821 | 34,840 | 114,981 |
| Coronary heart disease | CARDIoGRAMplusC4D^3^ | Males and females | Mostly European | 184,305 | 60,801 | 123,504 |
| Gout | Global Urate Genetics Consortium^4^ | Males and females | European | 69,374 | 2,115 | 67,259 |
| LDL cholesterol (mean) | Neale *et al* (UK Biobank)^5^ | Males and females | White British | 343,621 | - | - |
| Random glucose (mean) | Neale *et al* (UK Biobank)^5^ | Males and females | White British | 314,916 | - | - |
| Urate (mean) | Neale *et al* (UK Biobank)^5^ | Males and females | White British | 343,836 | - | - |
| LDL cholesterol (variance) | Lyon *et al* (UK Biobank)^6^ | Males and females | White British | 320,678 | - | - |
| Random glucose (variance) | Lyon *et al* (UK Biobank)^6^ | Males and females | White British | 291,579 | - | - |
| Urate (variance) | Lyon *et al* (UK Biobank)^6^ | Males and females | White British | 320,848 | - | - |

MRC-IEU, Medical Research Council Integrative Epidemiology Unit. LDL, low-density lipoprotein cholesterol.
